# Preventive dental visits and health literacy in patients with diabetes: a nationwide cross-sectional study

**DOI:** 10.1101/2024.07.01.24309770

**Authors:** Kyoko Saito, Yuki Kawai, Hirono Ishikawa, Takahiro Tabuchi, Keisuke Kuwahara

## Abstract

**Aim:** This cross-sectional study examined the association between health literacy and preventive dental visits in patients with diabetes.

**Methods:** We used cross-sectional data from the Japan COVID-19 and Society Internet Survey (JACSIS), a web-based nationwide survey. The participants were 1,441 patients reporting to have diabetes in 2020. Health literacy was measured using the validated scales for health literacy. Preventive dental visits in the past 12 months were self-reported. We estimated the multivariable-adjusted prevalence ratio (PR) for preventive dental visits.

**Results:** Over 50% of the patients had preventive dental visits in the past 12 months, and approximately one-third had high health literacy. Compared with the low health literacy group, the high health literacy group was more likely to engage in preventive dental visits (the multivariable-adjusted PR associated with high health literacy: 1.12 [95% confidence interval: 1.01 to 1.23]). Similar results were obtained when health literacy was treated as a continuous variable.

**Conclusions:** The present data from the JACSIS showed that health literacy was positively associated with preventive dental visits among patients with diabetes.

## 1. Introduction

Diabetes is a growing social and public health issue globally (1) including Japan (2). Recent statistics estimated that 537 million people worldwide (1) and 10 million in Japan (2) have diabetes. In patients, poor oral health, as indicated by periodontal disease, has been associated with an increased risk of diabetic complications, including cardiovascular complications, retinopathy, end-stage renal disease, and even deaths (3). Therefore, the development of effective strategies to prevent periodontal disease is needed for patients with diabetes.

Both self-care and professional care are important for the prevention of periodontal diseases (4). Therefore, regular dental visits are recommended for patients, particularly those with diabetes (4). For example, the Centers for Disease Control and Prevention (5) recommend getting a dental examinations at least once a year for patients with diabetes. The American Diabetes Association (6) also recommends comprehensive dental and periodontal examination for initial care management. However, the rate of dental visits among patients with diabetes remains low (e.g., 43% in Japan (7) and 61% in the US (8)), which are lower than those of patients without diabetes (8). This suggests an urgent need to identify modifiable factors that may promote dental visits among patients with diabetes.

Recently, health literacy has gained much attention in promoting behavioral changes (9). According to the World Health Organization (WHO), health literacy is defined as cognitive and social skills that determine the motivation and ability of individuals to gain access to, understand and use information in ways that promote and maintain good health’. Based on this definition, Nutbeam (10) proposed a health literacy model that includes three levels and assumes both individual and population benefits at each level: (i) functional/basic literacy, (ii) communicative/interactive literacy, and (iii) critical literacy.

The existing literature has shown that among patients with diabetes, health literacy is associated with self-care behaviors for diabetes (11), glycemic control (12) and retinopathy (13). Thus, health literacy may be associated with dental visits in patients with diabetes. To date, only one cross-sectional study has examined this association (14). Furthermore, although a preventive approach is important in dentistry (4; 5; 15), the previous study (14) did not distinguish preventive dental visits from traditional curative approaches. In addition, validity of measuring health literacy was unclear in that study (14). Therefore, we examined the association between health literacy and preventive dental visits using a validated questionnaire for health literacy among patients with diabetes in Japan.

## 2. Methods

### 2.1. Study design and data source

This cross-sectional study analyzed the baseline data of the Japan “COVID-19 and Society” Internet Survey (JACSIS), a web-based, self-reported questionnaire survey (16; 17). A total of 224,389 panelists aged 15 to 79 years from an internet research agency in Japan (Rakuten Insight, Inc.) were selected from approximately 2.2 million panelists, and asked about their willingness to participate in the survey. Eligible participants were selected using random sampling stratified by age, sex, and prefecture to represent the Japanese demographic composition. Questionnaires were distributed from August 25, 2020 to September 30, 2020; 28,000 respondents aged 15-79 years participated in this survey.

### 2.2. Participants

Of the 28,000 participants, we sequentially excluded 2,518 participants who provided invalid responses or met other exclusion criteria as with previous studies (17; 18); first, we excluded 1,955 participants providing invalid responses, using a dummy item stating: “Please choose the second option from the bottom.” We then excluded 422 participants who provided artificial or unnatural responses, defined as the use of recreational substances and medications, including sleeping pills, legal opioids, illegal opioids, organic solvents, designer drug, cannabis, cocaine. Lastly, we excluded 141 participants having all listed chronic diseases, including cancer, ischemic heart disease, stroke, diabetes, asthma, mental disorders, and so on. We additionally excluded one participant who reported an implausible body height (<100 cm). Of the remaining 25,481 participants (12,808 women and 12,673 men) aged 15-79 years (mean 48.8 years), we extracted the data of 1,441 participants aged 40-79 years who reported currently having diabetes. We restricted the participants to those aged 40 years or older because measurement of blood glucose levels and hemoglobin A1c levels are mandatory in this age group in the annual health checkups (19). We expected that the validity of self-reports on diabetes in this group would be better than that in younger groups. For sensitivity analyses, we additionally included 258 adults who reported to have diabetes in the past but not at the time of the study. As diabetes is not a curable disease, we expected that the participant characteristics would be somewhat different between those who reported having diabetes currently and those who had diabetes in the past but not currently.

### 2.3. Measurement of health literacy

Health literacy was measured using the Communicative and Critical Health Literacy (CCHL) scale which has been validated in Japanese (20). This scale measures communicative and critical health literacy, and comprised three items for communicative health literacy (items i–iii) and two items for critical health literacy (items iv–v). These items asked whether the participant were able to (i) collect health-related information from various sources, (ii) extract the information he wanted, (iii) understand and communicate the obtained information, (iv) consider the credibility of the information and (v) make decisions based on the information, specifically in the context of health-related issues (20). Each item is assessed using a five-point Likert scale ranging from 1 (strongly disagree) to 5 (strongly agree). We then calculated the total health literacy score by summing the score of each of the five responses divided by five, yielding scores of 1-5, where higher scores indicate greater health literacy. We reclassified the participants scoring less than 4 points as those with “low health literacy,” and participants scoring 4-5 points as those with “high health literacy” (20).

### 2.4. Measurement of past one years’ preventive dental visits

To measure preventive dental visits, respondents were asked, “In the past year, have you visited a dental clinic?” Dental visits included periodic checkups, dental cleaning, and tartar removal, but excluded visits for the treatment of cavities. Response options were “Yes” or “No.” The questionnaire on preventive dental visits was similar to those used in previous studies (21–23).

### 2.5. Other Covariates

The participants reported their socio-demographic factors such as age, sex, educational attainment, marital status, living arrangements, employment status, and annual household income in the past year (17) as well as health-related data, such as personal history of diseases, self-rated health, and body weight and height. History of diabetes was reported through four response options: none, having diabetes in the past but not currently, having diabetes currently with receiving treatment, having diabetes currently without treatment. In Japan, self-reported diabetes has been validated by using a similar question (24). Self-rated health was measured using 5-point scale (25; 26) and participants were classified into two groups: good (1-3 points) and poor (4-5 points) (25; 26). Body mass index (BMI; kg/m^2^) was calculated using body weight and height. BMI was reclassified as underweight (<18.5 kg/m^2^), normal weight (18.5 to <25.0 kg/m^2^), and overweight (≥25.0 kg/m^2^).

### 2.6. Ethics statement

As described previously (17), the present study was conducted in accordance with the ethical standards of the 1975 Declaration of Helsinki, as revised in 2013. Written informed consent was obtained from all participants before they responded to the online questionnaire. The study protocol, including the informed consent procedure, was approved by the ethics committees of the Osaka International Cancer Institute (no. 20084) and Teikyo University (no. 20-148-3). The Internet Survey Agency complies with the Act on the Protection of Personal Information in Japan. The data were anonymized; thus, the researchers did not have access to information that could identify individual participants. The participants were provided a credit point as an incentive for participation in this study.

### 2.7. Statistical analysis

Participant characteristics according to preventive dental visits are shown as numbers (%). We used Poisson regression to estimate multivariable-adjusted prevalence ratios (PRs) and the 95% confidence intervals (CIs) of preventive dental visits based on health literacy. We adjusted for age (years, continuous), sex, education attained (junior high school, high school, vocational school/technical college/junior college, university or higher, and others), marital status (married, unmarried, divorced, and widowed), living arrangement (living alone and living with someone), working status (currently working, unemployed or students, and retired), annual household income in 2019 (<3 million yen, 3 to <5 million yen, 5 to <7 million yen, ≥7 million yen, unclear, and do not want to answer), self-rated health status (good and poor), and BMI (<18.5, 18.5 to <25.0, and ≥25.0 kg/m^2^). Health literacy was treated as a dichotomous variable (low and high) and as a continuous variable. We also performed two sensitivity analyses. First, we conducted a stratified analysis by age (<65 years and ≥65 years) as older adults would have more dental or periodontal problems. Second, we conducted the sensitivity analyses by changing the definition of having diabetes as having diabetes currently or in the past. All analyses were performed using R4.1.3. or Stata 18.0. Statistical significance was set at p<0.05 (two-sided).

## 3. Results

Of the 1,441 participants who reported currently having diabetes, more than half had gone for preventive dental visits (n = 773, 53.6%). The average scores of health literacy was 3.5 (standard deviation: 0.7); approximately one in three patients (n = 508, 35.3%) had high health literacy. Table 1 shows the characteristics of the participants according to the number of preventive dental visits. Those who had preventive dental visits were slightly older, married or living with someone, likely to have a higher education and higher income, and not engaged in any work. They were also likely to report good self-rated health status and tended to have normal weight.

**Table 1.** Participant characteristics according to preventive dental visits among 1,441 diabetic patients.

|  | Preventive dental visits |  |
| --- | --- | --- |
|  | No | Yes |
| **N** | 668 | 773 |
| **Sex** |  |  |
| Male | 495 (74.1) | 583 (75.4) |
| Female | 173 (25.9) | 190 (24.6) |
| **Age, years** | 62.0±10.5 | 64.9±10.3 |
| **Education** |  |  |
| Junior high school | 48 (7.2) | 38 (4.9) |
| High school | 256 (38.3) | 251 (32.5) |
| Vocational school, technical college, or junior college | 93 (13.9) | 103 (13.3) |
| University or higher | 270 (40.4) | 379 (49.0) |
| Others | 0 (0) | 2 (0.2) |
| **Marital status** |  |  |
| Not married (unmarried, divorce, widowed) | 211 (31.6) | 176 (22.8) |
| Married | 457 (68.4) | 597 (77.2) |
| **Living arrangement** |  |  |
| Living alone | 127 (19.0) | 119 (15.4) |
| Living with someone | 541 (81.0) | 654 (84.6) |
| **Working status** |  |  |
| Currently working | 375 (56.1) | 384 (49.7) |
| Unemployed or students | 235 (35.2) | 284 (36.7) |
| Retired | 58 (8.7) | 105 (13.6) |
| **Annual household income last year** |  |  |
| <3 million yen | 155 (23.2) | 161 (20.8) |
| 3 million to <5 million yen | 139 (20.8) | 198 (25.6) |
| 5 to <7 million yen | 96 (14.4) | 102 (13.2) |
| ≥7 million yen | 142 (21.3) | 180 (23.3) |
| Unclear or do not want to answer | 136 (20.4) | 132 (17.1) |
| **Self-rated health** |  |  |
| Poor | 210 (31.4) | 190 (24.6) |
| Good | 458 (69.6) | 583 (75.4) |
| **Body mass index** |  |  |
| <18.5 kg/m<sup>2</sup> | 23 (3.4) | 26 (3.4) |
| 18.5 to <25.0 kg/m<sup>2</sup> | 336 (50.3) | 440 (55.9) |
| ≥25.0 kg/m<sup>2</sup> | 309 (46.3) | 307 (39.7) |
| Health literacy, scores | 3.4 ± 0.7 | 3.5 ± 0.7 |

Data are shown as n (%) or mean ± standard deviation).

Table 2 shows the relationship between preventive dental visits and health literacy.

**Table 2.** Prevalence ratios and 95% confidence intervals of preventive dental visits by health literacy among diabetic patients.

|  | N | Events | Unadjusted model |  | Adjusted model<sup>*</sup> |  |
| --- | --- | --- | --- | --- | --- | --- |
|  |  |  | PR (95% CI) | P value | PR (95% CI) | P value |
| **All participants** |  |  |  |  |  |  |
| Low (<4 points) | 933 | 470 (50.4) | Reference |  | Reference |  |
| High (≥ 4 points) | 508 | 303 (59.7) | 1.18 (1.08 to 1.30) | 0.001 | 1.12 (1.01 to 1.23) | 0.027 |
| 1 point increase (continuous) |  |  | 1.13 (1.05 to 1.22) | 0.001 | 1.09 (1.01 to 1.17) | 0.028 |
| **Aged <65 years** |  |  |  |  |  |  |
| Low (<4 points) | 357 | 183 (42.5) | Reference |  | Reference |  |
| High (≥ 4 points) | 304 | 121 (52.6) | 1.24 (1.05 to 1.46) | 0.011 | 1.19 (1.01 to 1.41) | 0.043 |
| 1 point increase (continuous) |  |  | 1.11 (0.99 to 1.25) | 0.085 | 1.07 (0.95 to 1.20) | 0.29 |
| **Aged ≥65 years** |  |  |  |  |  |  |
| Low (<4 points) | 502 | 287 (57.2) | Reference |  | Reference |  |
| High ($\geq 4$ points) | 278 | 182 (65.5) | 1.15 (1.02 to 1.28) | 0.02 | 1.09 (0.97 to 1.22) | 0.15 |
| 1 point increase (continuous)<sup>†</sup> |  |  | 1.14 (1.04 to 1.24) | 0.006 | 1.10 (1.00 to 1.21) | 0.045 |

Abbreviations: CI, confidence interval; PR, prevalence ratio.
\* Adjusted for sex, age, education, marital status, living arrangement, working status, annual household income, self-rated health, and body-mass index.

Participants with high health literacy were more likely to have had a preventive dental visit as compared to those with low health literacy; crude PR of preventive dental visit was 1.18 (95% CI: 1.08 to 1.30) for high health literacy group compared with low health literacy group. This relationship was slightly attenuated, but remained significant after additional adjustment for several covariates (aPR=1.12, 95% CI: 1.01, 1.23). A 1-point increase in health literacy was also significantly associated with preventive dental visits (aPR = 1.09). Analysis stratified by age groups showed similar point estimates, although some results were not statistically significant. As shown in Supplementary Tables 1, after enrolling 258 participants who reported having diabetes in the past but not currently, the associations were slightly weakened and some results became non-significant.

## **4.** Discussion

### 4.1. Discussion

In this cross-sectional study, health literacy was positively associated with preventive dental visits among patients with diabetes in Japan. The sensitivity analyses supported the main results. To the best of our knowledge, this is the first study to examine the relationship between health literacy and preventive dental visits in patients with diabetes.

Although there is no directly comparable study, our findings of significant and positive associations between health literacy and preventive dental visits are in line with at cross-sectional study by Rafferty et al (14). This study showed that individuals with low health literacy had lower adjusted odds ratios for dental visits; however, it did not distinguish between dental visits for preventive and treatment purposes, neither did it use any validated scales to assess health literacy. The present study using validated scales for health, underscores the importance of health literacy for patients with diabetes in promoting dental visits for preventive purposes.

### 4.2. Strengths and limitations

The strengths of this study include the large-scale sample of more than 1,000 patients with diabetes, use of a validated health literacy scale, and adjustment for many potential confounders. However, the results of this study should be interpreted in light of the following limitations. First, because of its cross-sectional design, it was not possible to confirm causality. Second, because we did not use objective measurements of glycemic conditions, patients with undiagnosed diabetes were not included in our analyses. Third, the validity of self-reported preventive dental visits was unclear, though previous studies have used similar self-reported questionnaires on dental visits (27; 28). Fourth, since the participants were recruited through an online survey, they were more likely to have better health literacy than the general populations. However, the CCHL scores in our study were not significantly different from those in previous studies (29), in which participants were recruited via mails. Fifth, although we adjusted for many potential confounders, unmeasured confounding factors including access to medical care may have affected the results. Finally, all participants were Japanese patients with diabetes. Therefore, caution should be exercised when generalizing the present findings to populations from other countries.

4.1. Conclusion

We found that health literacy was significantly associated with preventive dental visits among patients with diabetes in Japan. Healthcare professionals in diabetes care should consider patients’ health literacy levels to promote preventive dental visits. Further research is needed to confirm the present results using a longitudinal design and to determine whether health literacy education can promote preventive dental visits and prevent serious complications among patients with diabetes.

## Acknowledgement

The authors are thankful to those who involved in JACSIS.

## Data Availability

The data used in the present are not deposited in a public repository due to the containing of personally identifiable or potentially sensitive information. In accordance with the ethical guidelines’ regulations in Japan, dissemination of the data is restricted by the Research Ethics Committee of the Osaka International Cancer Institute. Any inquiries regarding the data use should go to Dr Takahiro Tabuchi. More details of data availability can be found on the website of JACSIS (https://jacsis-study.jp/howtouse/).

## References

1. International Diabetes Federation. IDF Diabetes Atlas, 10th edn. Brussels: International Diabetes Federation, 2021

2. Ministry of Health, Labour and Welfare. *The National Health and Nutrition Survey in Japan,* 2016. Tokyo: Ministry of Health, Labour and Welfare.

3. Casanova L, Hughes FJ, Preshaw PM. Diabetes and periodontal disease: a two-way relationship. Br Dent J 2014;217:433–437

4. Office of Disease Prevention and Health Promotion, Department of Health and Human Service. Healthy People 2030: Oral Conditions. Available from https://health.gov/healthypeople/objectives-and-data/browse-objectives/oral-conditions. Accessed 01-24 2024

5. Centers for Disease Control and Prevention. How to Promote Oral Health for People With Diabetes, Available from https://www.cdc.gov/diabetes/professional-info/health-care-pro/diabetes-oral-health.html. Accessed 01-24 2024

6. American Diabetes Association. Oral Health. Available from https://diabetes.org/health-wellness/keeping-your-mouth-healthy. Accessed 01-24 2024

7. Inagaki K, Kikuchi T, Noguchi T, Mitani A, Naruse K, Matsubara T, Kawanami M, Negishi J, Furuichi Y, Nemoto E, Yamada S, Yoshie H, Tabeta K, Tomita S, Saito A, Katagiri S, Izumi Y, Nitta H, Iwata T, Numabe Y, Yamamoto M, Yoshinari N, Fujita T, Kurihara H, Nishimura F, Nagata T, Yumoto H, Naito T, Noguchi K, Ito K, Murakami S, Nishimura R, Tajima N. A large-scale observational study to investigate the current status of diabetic complications and their prevention in Japan (JDCP study 6): baseline dental and oral findings. Diabetol Int 2021;12:52–61

8. Luo H, Bell RA, Wright W, Wu Q, Wu B. Trends in annual dental visits among US dentate adults with and without self-reported diabetes and prediabetes, 2004-2014. J Am Dent Assoc 2018;149:460-469

9. Sørensen K, Van den Broucke S, Fullam J, Doyle G, Pelikan J, Slonska Z, Brand H, European H-ECHLP. Health literacy and public health: a systematic review and integration of definitions and models. BMC Public Health 2012;12:80

10. Nutbeam D. Promoting health and preventing disease: an international perspective on youth health promotion. J Adolesc Health 1997;20:396–402

11. Ong-Artborirak P, Seangpraw K, Boonyathee S, Auttama N, Winaiprasert P. Health literacy, self-efficacy, self-care behaviors, and glycemic control among older adults with type 2 diabetes mellitus: a cross-sectional study in Thai communities. BMC Geriatr 2023;23:297

12. Alvarez PM, Young LA, Mitchell M, Blakeney TG, Buse JB, Vu MB, Weaver MA, Rees J, Grimm K, Donahue KE. Health Literacy, Glycemic Control, and Physician-Advised Glucose Self-Monitoring Use in Type 2 Diabetes. Diabetes Spectr 2018;31:344–347

13. Schillinger D, Grumbach K, Piette J, Wang F, Osmond D, Daher C, Palacios J, Sullivan GD, Bindman AB. Association of health literacy with diabetes outcomes. JAMA 2002;288:475–482

14. Rafferty AP, Winterbauer NL, Luo H, Bell RA, Little NRG. Diabetes Self-Care and Clinical Care Among Adults With Low Health Literacy. J Public Health Manag Pract 2021;27:144–153

15. World Health Organization. Oral health. https://www.who.int/news-room/fact-sheets/detail/oral-health [accessed 2024-01-24].

16. Matsuyama Y, Aida J, Takeuchi K, Koyama S, Tabuchi T. Dental Pain and Worsened Socioeconomic Conditions Due to the COVID-19 Pandemic. J Dent Res 2021;100:591–598

17. Kuwahara K, Kato M, Ishikawa H, Shinozaki T, Tabuchi T. Association between watching wide show as a reliable COVID-19 information source and preventive behaviors: A nationwide survey in Japan. PLOS ONE 2023;18:e0284371

18. Miyawaki A, Tabuchi T, Tomata Y, Tsugawa Y. Association between participation in the government subsidy programme for domestic travel and symptoms indicative of COVID-19 infection in Japan: cross-sectional study. BMJ Open 2021;11:e049069

19. Ministry of Health, Labour and Welfare. “Specific Health Checkups and Specific Health Guidance”. (in Japanese) Available from https://www.mhlw.go.jp/stf/seisakunitsuite/bunya/0000161103.html. Accessed 1-24 2024

20. Ishikawa H, Nomura K, Sato M, Yano E. Developing a measure of communicative and critical health literacy: a pilot study of Japanese office workers. Health Promot Int 2008;23:269–274

21. Foiles Sifuentes AM, Castaneda-Avila MA, Lapane KL. The relationship of aging, complete tooth loss, and having a dental visit in the last 12 months. Clin Exp Dent Res 2020;6:550–557

22. Spinler K, Aarabi G, Walther C, Valdez R, Heydecke G, Buczak-Stec E, König HH, Hajek A. Determinants of dental treatment avoidance: findings from a nationally representative study. Aging Clin Exp Res 2021;33:1337–1343

23. Zhang Y, Leveille SG, Shi L, Camhi SM. Disparities in Preventive Oral Health Care and Periodontal Health Among Adults With Diabetes. Prev Chronic Dis 2021;18:E47

24. Goto A, Morita A, Goto M, Sasaki S, Miyachi M, Aiba N, Kato M, Terauchi Y, Noda M, Watanabe S, Saku Cohort Study G. Validity of diabetes self-reports in the Saku diabetes study. J Epidemiol 2013;23:295–300

25. Kawachi I, Kennedy BP, Glass R. Social capital and self-rated health: a contextual analysis. Am J Public Health 1999;89:1187–1193

26. Sugisawa H, Harada K, Sugihara Y, Yanagisawa S, Shinmei M. Socioeconomic status and self-rated health of Japanese people, based on age, cohort, and period. Popul Health Metr 2016;14:27

27. Sakalauskiene Z, Maciulskiene V, Vehkalahti MM, Kubilius R, Murtomaa H. Characteristics of dental attendance among Lithuanian middle-aged university employees. Medicina (Kaunas) 2009;45:312-319

28. Kosteniuk J, D’ Arcy C. Dental service use and its correlates in a dentate population: an analysis of the Saskatchewan population health and dynamics survey, 1999-2000. J Can Dent Assoc 2006;72:731

29. Kimura N, Kobayashi T. [Association of health literacy with hypertension, diabetes, and dyslipidemia: A cross-sectional survey of a regional Japanese community]. Nihon Koshu Eisei Zasshi 2020;67:871–880

