## Supplementary Table for "Preventive dental visits and health literacy in patients with diabetes: a nationwide cross-sectional study"

**Supplementary Table 1. Prevalence ratios and 95% confidence intervals of preventive dental visits and health literacy after including participants who reported to have diabetes in the past but not currently**

| | N | Events | Unadjusted model | | Adjusted model* | |
|---|---|---|---|---|---|---|
| | | | PR (95% CI) | P value | PR (95% CI) | P value |
| **All participants** | | | | | | |
| Low (<4 points) | 1,110 | 557 (50.2) | Reference | | Reference | |
| High (≥ 4 points) | 589 | 341 (57.9) | 1.15 (1.05 to 1.27) | 0.002 | 1.09 (0.99 to 1.19) | 0.064 |
| 1 point increase (continuous) | | | 1.11 (1.05 to 1.20) | 0.001 | 1.07 (1.00 to 1.15) | 0.047 |
| **Aged <65 years** | | | | | | |
| Low (<4 points) | 513 | 216 (42.1) | Reference | | Reference | |
| High (≥ 4 points) | 273 | 142 (52.0) | 1.24 (1.06 to 1.44) | 0.007 | 1.17 (1.00 to 1.36) | 0.055 |
| 1 point increase (continuous) | | | 1.13 (1.01 to 1.26) | 0.029 | 1.09 (0.97 to 1.22) | 0.15 |
| **Aged ≥65 years** | | | | | | |
| Low (<4 points) | 597 | 341 (57.1) | Reference | | Reference | |
| High (≥ 4 points) | 316 | 199 (63.0) | 1.10 (0.99 to 1.23) | 0.081 | 1.06 (0.95 to 1.19) | 0.27 |
| 1 point increase (continuous) | | | 1.09 (1.00 to 1.19) | 0.039 | 1.06 (0.97 to 1.16) | 0.17 |

Abbreviations: CI, confidence interval; PR, prevalence ratio.

*Adjusted for sex, age, education, marital status, living arrangement, working status, annual household income, self-rated health, and body-mass index.
